# The hidden hand of asymptomatic infection hinders control of neglected tropical diseases

**DOI:** 10.1101/2023.10.02.23296422

**Authors:** Kat S. Rock, Lloyd A.C. Chapman, Andrew P. Dobson, Emily R. Adams, T. Déirdre Hollingsworth

**Author notes:** Equal contributors (Kat S. Rock, Lloyd A.C. Chapman). Equal contributors (Andrew P. Dobson, Emily R. Adams, T. Déirdre Hollingsworth). **Author contributions** Conceptualization: TDH, KSR, LACC, APD, ERA, Methodology: KSR, LACC, APD, TDH, Software: KSR, Validation: LACC, Formal Analysis: KSR, LACC. Investigation: KSR, LACC, TDH, Resources: KSR, LACC, TDH, Data Curation: LACC, KSR, Writing – Original Draft Preparation: ERA, KSR, LACC, APD, Writing – Reviewing and Editing: KSR, LACC, APD, ERA, TDH, Visualisation: KSR, LACC, TDH, Supervision: ERA, TDH, Project administrations: ERA, TDH, Funding Acquisition: TDH. The authors declare no competing interests.

## Abstract

Neglected tropical diseases are responsible for considerable morbidity and mortality in low-income populations. International efforts have reduced their global burden, but transmission is persistent and case-finding-based interventions rarely target asymptomatic individuals. We develop a generic mathematical modelling framework for analysing the dynamics of visceral leishmaniasis in the Indian sub-continent (VL), *gambiense* sleeping sickness (gHAT), and Chagas disease and use it to assess the possible contribution of asymptomatics who later develop disease (pre-symptomatics) and those who do not (non-symptomatics) to the maintenance of infection. Plausible interventions, including active screening, vector control and reduced time to detection, are simulated for the three diseases. We conclude that currently available tools could bring new infections under control for gHAT and Chagas disease – albeit over a long period of time for Chagas disease – but that VL will be more difficult to control with existing interventions. gHAT protocols that allow for treatment of parasitologically positive asymptomatics result in better control of infection compared to syndromic diagnosis required for VL. This work highlights that the high asymptomatic contribution to transmission for Chagas and gHAT and the apparently high basic reproductive number of VL may greatly undermine long-term control, but that precise predictions are hampered by uncertainty in natural history.

## Introduction

Neglected tropical diseases (NTDs) have a major impact on human health and economic development [1]. A substantial component of this is the high proportion of asymptomatically infected individuals who may transmit infection while not developing, or only slowly developing, symptoms that qualify them for treatment [2]. A cruel irony is that treatment can be more effective when patients are asymptomatically infected, as has been shown for Chagas disease in the Americas [3-5]. Here we focus on three major vector-borne NTDs on different continents: visceral leishmaniasis (VL) in the Indian subcontinent, *gambiense* human African trypanosomiasis (sleeping sickness, gHAT) and Chagas disease in the Americas. We use a parameterised mathematical model to estimate the relative contribution of asymptomatic individuals to onward transmission. Although intervention programmes exist for each disease, they typically do not address asymptomatic contributions to infection and this may appreciably undermine control gains [6-9]. We develop a general model that quantifies the impact of the transmission by asymptomatic hosts on the long-term sustainability of intervention programmes. The analysis illustrates the importance of tackling asymptomatic infection using better diagnostic tools, new treatment options, and vector control.

Many pathogens have long asymptomatic periods when transmission may occur and the host is unaware they are infected. COVID-19 and HIV are notable examples [10, 11], but several important vector-borne diseases also have these properties, notably VL, gHAT and Chagas disease [2]. These hidden reservoirs of infection present a challenge to public health, particularly if asymptomatic individuals are major contributors to transmission. Recognising and quantifying the magnitude of this problem is likely to lead to notable modification of current public health policies, demonstrating a need to: (i) more systematically diagnose and treat asymptomatic hosts, and (ii) reduce transmission from asymptomatic hosts using suitable interventions.

Our goal is to address three questions: (a) How prevalent are asymptomatic infections in VL, gHAT and Chagas disease and how infectious are they to vectors? (b) What is their relative contribution to transmission compared to hosts with overt symptoms? (c) How does a better understanding of the role of asymptomatic infections inform attempts to control and eliminate vector-borne NTDs and other similar, or emerging, pathogens?

### Epidemiology and control

VL, gHAT and Chagas disease are vector-borne parasitic diseases caused by flagellated protozoa belonging to the Class Kinetoplastida. They are the subject of broad-scale interventions (see Table S1) with targets for elimination as a public health problem (Chagas and VL) and elimination of transmission (gHAT) by the end of 2030 [12]. All three are considered fatal if left untreated in the symptomatic form, and all three have substantial and (largely) untreated asymptomatic populations with uncertain natural history [3, 13-15] – this has impeded previous analyses of their impact on transmission. One great challenge is that the definition of asymptomatic infection varies according to the diagnostic used and therefore changes over time [16, 17]. Here we have defined asymptomatic infection as either pre-symptomatic, for those that go on to develop disease, or non-symptomatic, for those that never develop symptoms (see disease-specific definitions in Table S2). We have collated the relatively limited available parasitological, epidemiological and xenodiagnostic data to quantify their natural history of disease (Data S1, Fig. 1B), which we combine with estimates of the proportion of infections in each group to estimate the potential contributions of each group to transmission (Fig. 1C).

**Figure 1.**
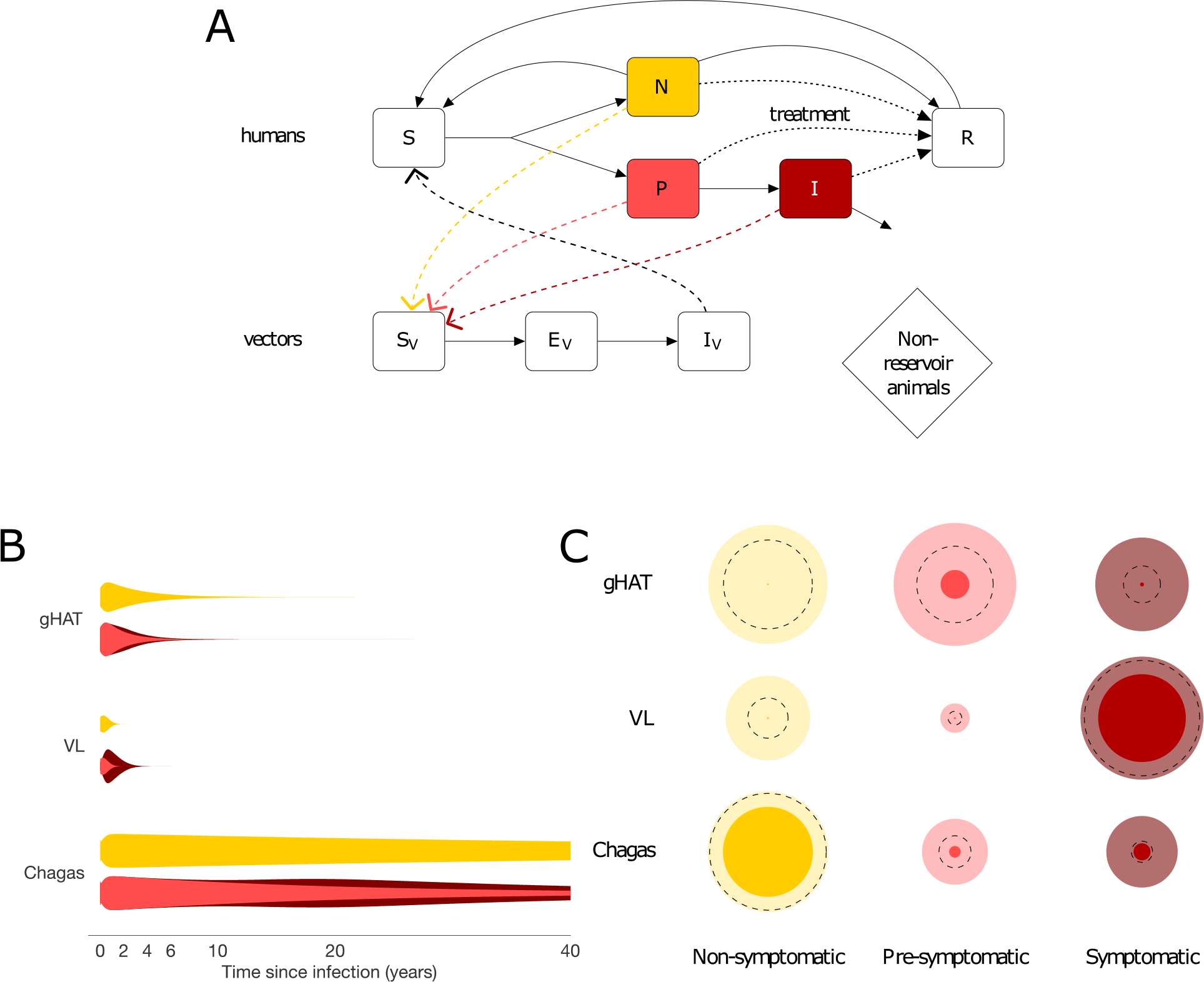
(A) Flow diagram for generic model of transmission of vector-borne kinetoplastid disease showing progression of susceptible individuals (*S*) to asymptomatic infection following infection by infectious vectors (*I*_V_) and infection of susceptible vectors (*S*_V_) by infectious individuals. Asymptomatics are divided into two groups who are usually indistinguishable by standard diagnostics, non-symptomatics (*N*) and pre-symptomatics (*P*) who go on to develop symptoms (*I*). Disease progression shown by solid arrows, transmission by dashed arrows, and treatment by dotted arrows. (B) Schematic of relative infectiousness (represented by width of violin plots) of non-symptomatic, pre-symptomatic and symptomatic infection over time since infection for *gambiense* human African trypanosomiasis (gHAT), visceral leishmaniasis (VL) in the Indian sub-continent and Chagas disease. Schematics derived from a handful of parasitological studies and limited xenodiagnostic studies (which assess transmission from infected individuals towards vectors) and epidemiological studies of the natural history of disease, alongside indications that symptoms are generally associated with higher parasite loads (Data S1). Uncertainty in durations is shown by thin tails. (C) Estimated relative contribution (represented by bubble sizes) to *R*_0_^2^ (secondary infections) of non-symptomatic, pre-symptomatic and symptomatic infection for the three diseases in the absence of interventions. Darker inner and lighter outer bubbles represent minimum and maximum possible contribution to *R*_0_^2^ based on lower and upper bounds for parameters from the literature. Dotted lines represent contributions based on mid-range, illustrative values of parameters from the literature (see Data S1 and SM).

#### Epidemiological metrics

Three key metrics are used throughout the present study to evaluate the potential impact of different interventions on each disease and compare and contrast the different diseases:

- the full-cycle (host-to-host) basic reproduction number, *R*_0_^2^, for a vector-borne disease, defined as the number of secondary infections generated by a single infectious individual in an entirely susceptible population, and the full-cycle control reproduction number, *R*_c_^2^, the same quantity accounting for control interventions;
- the contribution of asymptomatics to *R*_0_^2^, denoted *θ*, and that specifically from pre-symptomatics *θ*_P_ and non-symptomatics *θ*_N_;
- the contribution of asymptomatics, and that of pre-symptomatics and non-symptomatics, to transmission over time.

#### gHAT

*Trypanosoma brucei gambiense* is transmitted by the bites of infected tsetse in sub-Saharan Africa. Due to robust intervention programmes, prevalence has recently been brought to low levels and fewer than 800 annual global cases were reported during 2020–2022 [18]. For gHAT most symptomatic infections present within a few years (Fig. 1B), although there are much longer examples [19]. There is evidence of trypanosomes circulating in skin and other organs [20] and one study shows that animals/mice with no detectable blood parasites can still infect tsetse through their skin at ∼58% of the frequency compared to those with blood infections [21]. The current drug options preclude treatment of serological suspects without visualisation of the parasite (even if symptomatic), but symptoms are not required to administer treatment. To date, analyses of gHAT transmission have not been able to quantify the extent to which asymptomatics hinder efforts to achieve elimination, although modelling increasingly suggests that systematically undetected human infections are driving transmission [17, 22-24].

In a fully susceptible population, the relative contribution of each type of infection to transmission is essentially the product of the proportion of infections it represents, the average duration of infection and its relative infectiousness to the vector. When calculating this quantity for gHAT, available data suggest that pre- and non-symptomatic individuals are contributing to a large proportion of transmissions (Fig. 1C, Data S1). Current screening and vector control activities reduce transmission by targeting all three groups, but imperfect diagnostic algorithm sensitivity (especially for skin infections [24]) and non-participation of individuals who do not feel sick in active screening [25] could hinder efficiency of the interventions targeted at humans. Vector control activities, primarily using Tiny Targets, have been shown to be very effective at reducing tsetse populations [26] but are not conducted in all endemic settings.

#### VL

*Leishmania donovani* is transmitted by female sandflies. Incidence of the disease in the Indian sub-continent (ISC), where there is an active elimination campaign, has decreased dramatically (falling from 37,000 cases in 2011 to 1,500 cases in 2021 [18]). Infection is characterised by a long pre-symptomatic period, and many individuals who are asymptomatically infected (Fig. 1A). Recent xenodiagnostic studies [27, 28] suggest that asymptomatically infected individuals are significantly less infectious to sandflies than symptomatic VL cases (with a probability of transmission of <2% compared to 67–78% for VL cases). Nevertheless, experimental data suggest that these studies may have limited sensitivity to detect transmission to sandflies [29] and outbreaks of VL in regions where there are asymptomatic individuals but virtually no VL cases have been reported suggest asymptomatic individuals can transmit to sandflies [30, 31]. Post kala-azar dermal leishmaniasis (PKDL), a skin infection of Leishmania parasites which mostly occurs after treatment for VL, is an added challenge in controlling transmission of VL since PKDL cases constitute a potential large and untreated reservoir of infection, especially as incidence of VL decreases [32], as some cases are infectious to sandflies [27, 33]. Here, we do not consider PKDL explicitly, with the understanding that the inclusion of PKDL will only boost the role of non-VL infections and can be revisited as more data become available. Previous modelling analyses have suggested that asymptomatic individuals are likely to be far less infectious to sandflies than symptomatic cases (1/80th–1/40th as infectious), but may be a major contributor to overall disease transmission because they represent a large proportion of the infected population [34-36] particularly once immunity declines in the population post-outbreaks [37, 38]. Without addressing these putative infection reservoirs, the gains made during the elimination campaign may be compromised.

In contrast to gHAT, the currently available estimates suggest that symptomatic VL infection is the largest contributor to transmission, stressing the importance of effective case finding (Data S1, Fig. 1C). However, there is large uncertainty in the proportion of infections that progress to disease and in the relative infectiousness of different infection states. The main interventions against VL currently employed in the Indian subcontinent are early detection and treatment of symptomatic cases and indoor residual spraying of insecticide (IRS) aimed at reducing vector abundance [39, 40]. There is currently no treatment available for asymptomatic infection due to the high toxicity and potential side-effects associated with VL and PKDL treatment drugs [41].

#### Chagas disease

Chagas disease is endemic in South America and spreading into the United States, Europe and South East Asia following migration of asymptomatically infected human hosts. It is associated with poor housing since the triatomine bugs that transmit the disease live in cracks in the walls of rural mud houses. Bed bugs may act as vectors in areas where triatomines are controlled or absent [42]. There is a large reservoir of infection in wildlife, but much of the incidence is believed to be driven by a very large pool of asymptomatic human infection in areas of anthroponotic transmission [43]. Differentiation of asymptomatic infection and disease is essential since the period prior to symptom onset is typically several decades (Fig. 1B); the symptomatic stage is potentially fatal, even after several years of treatment. An additional challenge related to the asymptomatic nature of Chagas disease is the contamination of blood banks in endemic and non-endemic countries when donors do not know their infection status. Modelling has shown that identification and treatment of asymptomatics can substantially reduce the prevalence of Chagas and aid elimination prospects [7]. The relative contributions to transmission, derived from available literature (Data S1, Fig. 1C), clearly support the view that asymptomatic infection is a major contributor to transmission, even if it is only 33% as infectious as symptomatic infection, due to its long duration. Historically, vector control through IRS has been the main strategy used to reduce transmission of Chagas, but increasing availability of anti-trypanosomal drugs has enabled broader treatment, including for chronic infection in adults [3, 7].

For all three diseases, there is uncertainty in the relative contributions of non-symptomatics, pre-symptomatics and symptomatics due to the wide ranges of the parameter values in the literature (Data S1). This emphasises the need for better empirical data and diagnostics, since the contributions of the different states could be roughly similar, or differ by orders of magnitude.

Although there are marked differences in the epidemiology of these diseases, particularly the timescale for disease progression, they share important features in terms of the public health relevance of the role of asymptomatics and the need for quantitative investigation of their dynamics. The paucity of experimental data increases our dependence upon mathematical modelling to predict the impact of the asymptomatic population on interventions as we move towards the peri-elimination era. We group the characteristics of asymptomatic infection across the diseases to see if generalisable themes that are directly relevant to intervention measures arise from a cross-disease comparison using a generic transmission model (see Supplementary Text S1).

## Results

We build upon insights developed by Fraser et al. [44] and consider the role of interventions that differentially impact the asymptomatic pool: we first use a flexible, generic modelling framework (Fig. 1A) to consider the relationship between the basic reproduction number for a vector-borne disease, *R*_0_^2^, and ***θ***, the proportion of new infections arising from both non-symptomatic and pre-symptomatic individuals (Fig. 2). Reproduction numbers below one will eventually lead to elimination of infection, whereas values above this threshold mean new infections will continue. Broadly speaking, in the absence of effective interventions to combat non-symptomatic and pre-symptomatic infection (diseases in the upper right-hand corner with higher *R*_0_^2^ and higher ***θ***) are harder to control. Even in the presence of uncertainty in the natural history, our estimates suggest treatment solely of symptomatic individuals for gHAT is unlikely to interrupt transmission (Fig. 2B). Additional treatment of asymptomatics (beyond those targeted by current mass screening) is much more likely to lead to control (Fig. 2C). In contrast, it may be possible to control VL by treating 90% of symptomatic individuals if asymptomatic and pre-symptomatic infectiousness to sandflies is low. As most *T. cruzi* infections are caused by individuals with asymptomatic Chagas disease, treatment solely of symptomatics is unlikely to lead to a substantial reduction in new infections. The higher reproduction numbers for VL and Chagas mean that it is likely that a higher proportion of pre-symptomatic and/or non-symptomatic infections would need to be treated. Moderate levels of vector control appear sufficient to control gHAT but may not be sufficient on their own to control Chagas disease or VL (Fig. 2D). If treatment of symptomatic people is added to vector control it is more likely that transmission of Chagas disease and VL would be interrupted.

**Figure 2.**
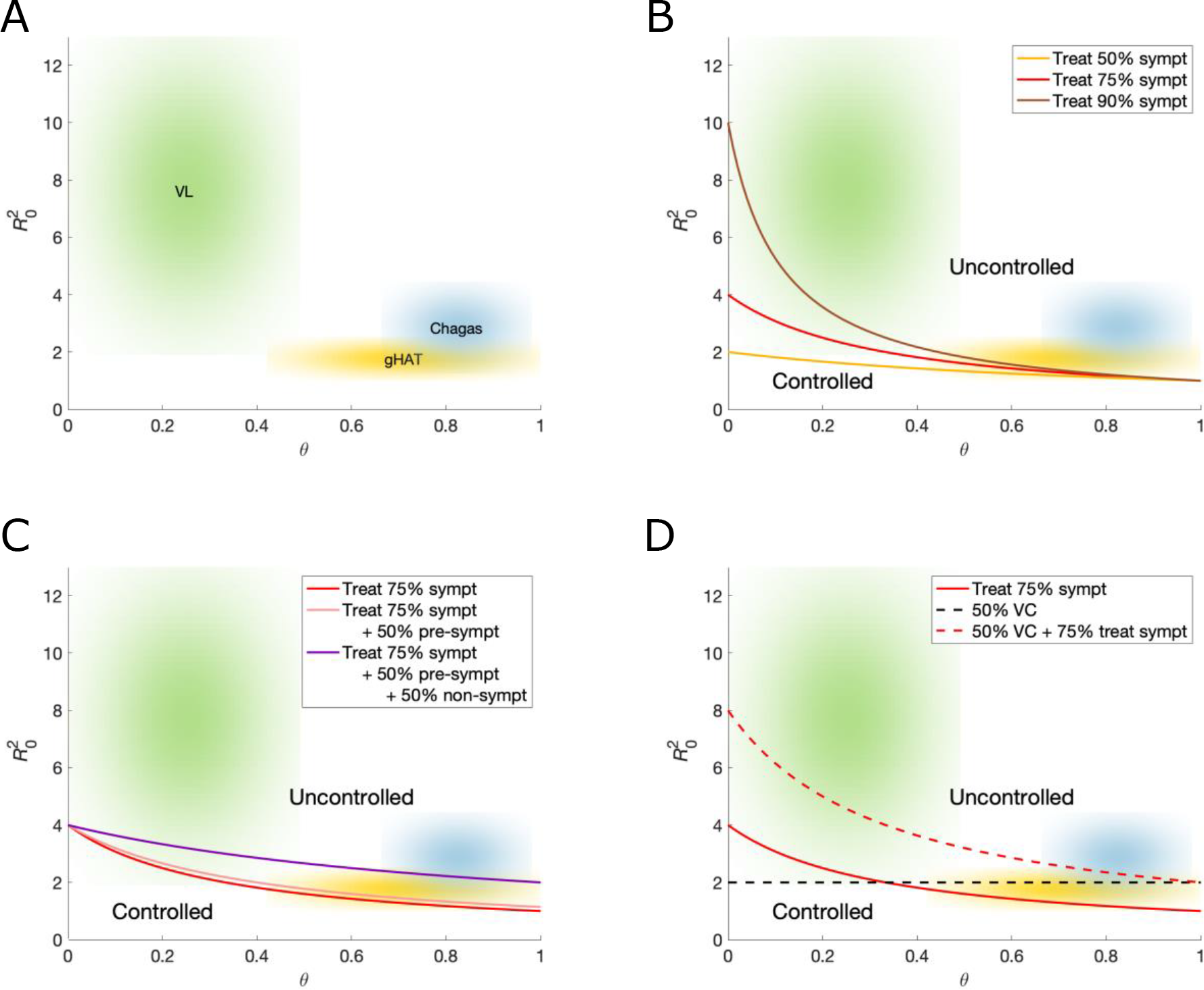
Potential for control of VL, gHAT and Chagas disease under different combinations of generic interventions. (A) Plot illustrating the relative position of each of VL (green), gHAT (yellow), and Chagas disease (blue) in the phase space determined by the full-cycle reproduction number of the disease, *R*_0_^2^ (the average number of secondary infections caused by one infectious individual in an entirely susceptible population), and the proportional contribution of non-symptomatic and pre-symptomatic individuals to *R*_0_^2^, ***θ***. Clouds represent parameter uncertainty. (B)–(D) Sensitivity of the diseases to different interventions. The curves delineate successful control (below the line) with each intervention: (B) treating symptomatic individuals; (C) treating symptomatic individuals and pre-symptomatic and/or non-symptomatic individuals; (D) vector control (VC) – achieving 50% vector control corresponding to halving the vector population, reducing the bite rate or shortening vector life expectancy.

Simulated interventions against gHAT and Chagas disease reduce transmission substantially, below the critical threshold for elimination, but the reduction in transmission takes a long period of time due to the long timescale of infection (especially for Chagas disease, Fig. 3A and C). Conversely, the strategy for VL is not sufficient for elimination, although transmission reduction happens quite swiftly (Fig. 3B). The contribution of transmission shifts towards asymptomatics when interventions target symptomatics, highlighting the importance of surveillance and treatment of all host groups to achieve elimination (see Fig. S3–5 for alternative intervention strategies).

**Figure 3.**
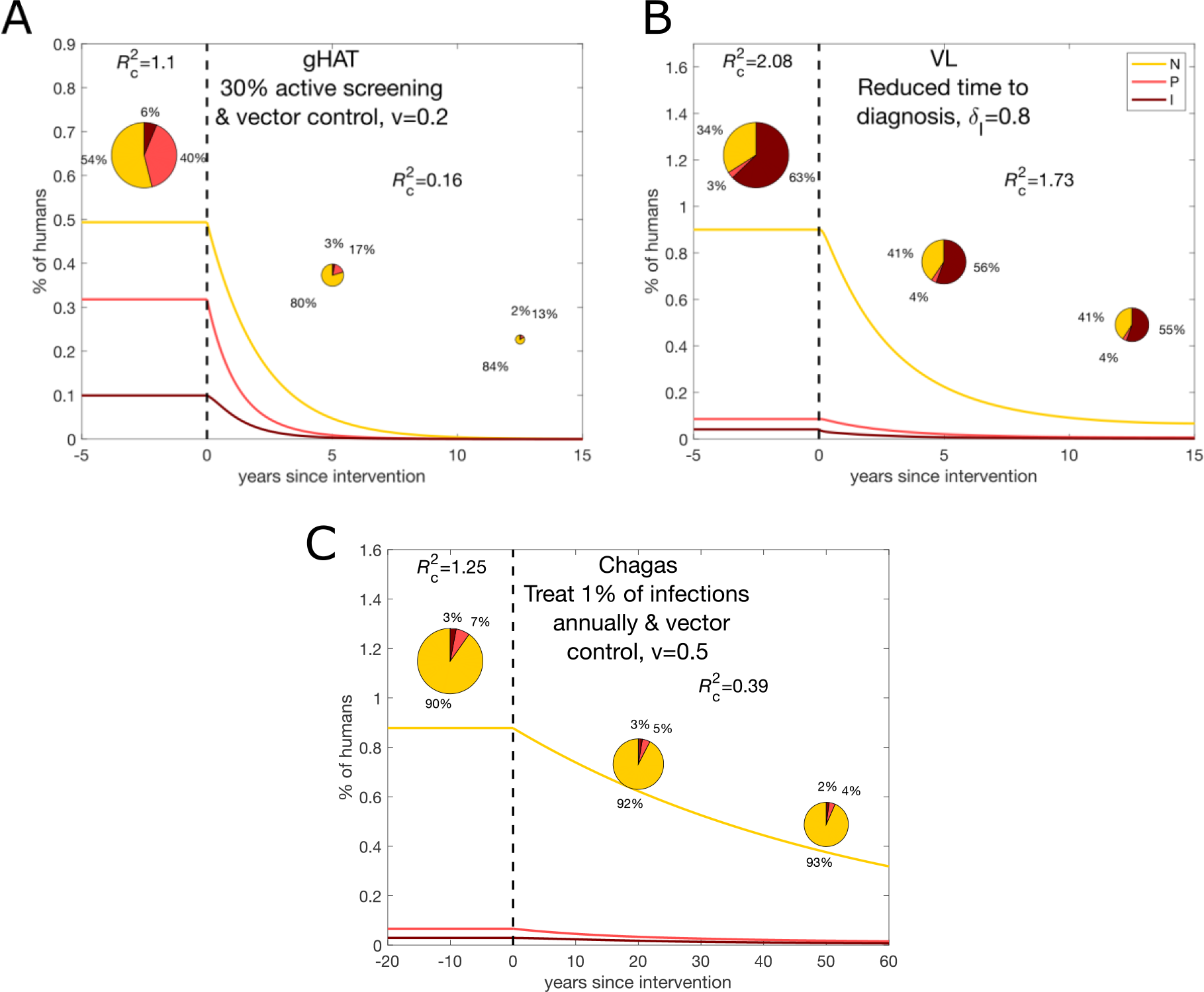
Change in the contribution of non-symptomatic, pre-symptomatic, and symptomatic individuals to transmission over time as interventions are carried out for realistic interventions for each of the three diseases. (A) gHAT. (B) VL. (C) Chagas disease. The lines show the proportions of the population in each infection state: non-symptomatic (*N*), pre-symptomatic (*P*) and symptomatic (*I*). The pie charts show the contribution of these groups to new infections at particular points in the timeline. *R*_c_^2^ denotes the full-cycle reproduction number accounting for control, 1-*v* denotes the proportional reduction in the vector-to-host ratio due to vector control (such that 1 indicates no vector control), and ***δ***_I_ denotes the proportion of symptomatic infections that are treated before death.

## Discussion

Our results suggest that gHAT may be the easiest of the three diseases to control with combinations of existing interventions. We predict a change from non-symptomatic and pre-symptomatic individuals being responsible for approximately 50% and 40% of overall transmission respectively, to being responsible for roughly 80% and 20% respectively after 5 years. It is important to note that combining interventions targeted at the vector and at asymptomatic individuals (active screening), leads to the target being reached more quickly.

For Chagas disease, it is widely known that treatment is more successful for asymptomatics and likely to control their parasite burden. It not only prevents disease progression but also reduces onward transmission in the community leading to a reduction in incidence. Changes in interventions that permit this will only be successful if better diagnostic techniques are developed and more widely used. In particular, diagnosis of asymptomatic hosts and treatment with novel drugs that slow pathogen development and reduce transmission from asymptomatics (rather than attempt to eliminate infection) may prove more viable than only treating overt infections.

The situation for VL is less clear due to limited data on the relative infectiousness of different infection states, the inability to treat asymptomatic cases with the currently available drugs, and the challenges of PKDL and HIV co-infection. The literature suggests that VL has a higher reproduction number than Chagas disease and gHAT, and that it is unlikely its transmission can be interrupted without multiple interventions to reduce the time to diagnosis, target the vector, and address the issues of PKDL and asymptomatic reservoirs of infection.

Key to addressing the issue of asymptomatic infection is understanding that infection dynamics are not in equilibrium and that the contribution of different groups to transmission is continually changing. Whilst it is not possible to capture all aspects of the natural histories and transmission cycles of all vector-borne NTDs in a single model, (e.g. possible animal transmission for Chagas and gHAT, PKDL for VL), the model presented here provides a general framework for considering the role of asymptomatic infection in transmission of vector-borne NTDs that can be built upon for specific diseases, particularly malaria and vector-borne emerging pathogens.

A limitation of all policy-relevant work to date on these diseases is the scarcity of experimental data on which to base assumptions. Our ability to design accurate models could greatly improve with more empirical data using (i) xenodiagnostic techniques where vectors take a bloodmeal from an asymptomatic individual with longitudinal follow-up, (ii) experimental animal infections where vectors take a blood meal from infected animals, and (iii) quantitative comparison with symptomatic patients. A particular challenge that contributes to the uncertainty in the definition of asymptomatic infection is finding good diagnostics for identifying asymptomatics and stratifying them into those who are likely to develop symptoms and those who are not [45-47]. Such diagnostics would enable better patient care and selection of interventions to reduce transmission [38, 48].

This work highlights the currently under-appreciated challenge and importance of asymptomatic infections in three of the world’s major NTDs. We have shown that there is potentially a major reservoir of infection in non-symptomatic and pre-symptomatic hosts, and that additional interventions targeted at these groups could lead to considerable progress in the elimination of Chagas, VL, gHAT and other pathogens. Not only would transmission in the community be reduced, but the burden on the public health system would be lessened if individuals did not progress to disease. Expanding public health programmes for these NTDs to focus on systematic testing and treatment of asymptomatic individuals requires a conceptual shift in the underlying control approach, similar to that that occurred during the COVID-19 pandemic, to develop new effective pathways to combat these diseases in which asymptomatic hosts make large contributions to transmission.

## Data Availability

Data is available as supplementary information, and code available on request

## Acknowledgements

All authors gratefully acknowledge funding of the NTD Modelling Consortium by the Bill and Melinda Gates Foundation OPP1053230 (in partnership with the Task Force for Global Health), OPP1184344, OPP1177824 and INV-005121. This supplement is sponsored by funding of Professor T. Déirdre Hollingsworth’s research by the Li Ka Shing Foundation at the Big Data Institute, Li Ka Shing Centre for Health Information and Discovery, University of Oxford and funding of the NTD Modelling Consortium by the Bill & Melinda Gates Foundation (INV-030046). The views, opinions, assumptions or any other information set out in this article are solely those of the authors and should not be attributed to the funders or any person connected with the funders. The funders had no role in study design, data collection and analysis, decision to publish, or preparation of the manuscript.

