## Supplementary information for "The hidden hand of asymptomatic infection hinders control of neglected tropical diseases"

#### Summary of available interventions

Table S1 gives a synopsis of various interventions that are current in use in (some) endemic settings aimed at controlling and/or eliminating *gambiense* human African trypanosomiasis (gHAT), visceral leishmaniasis (VL) in the Indian subcontinent, and Chagas disease. Interventions for these disease can often vary considerable by region, depending on disease incidence, available financial and personnel resources to conduct the interventions, and geographical accessibility or suitability. We have not included tools in the pipeline, but the availability of emerging tools could expand intervention options; e.g. for gHAT a new, single-dose oral drug – acoziborole – will hopefully gain approval for use in the near future and, if the safety profile allows, could enable treatment based on serological diagnosis alone, compared to current need to have parasitological confirmation prior to treatment [14].

Table S1: High-level overview of currently available interventions against gHAT, VL and Chagas disease

| | gHAT | VL | Chagas |
|---|---|---|---|
| Passive detection/screening | Self-presentation of symptomatic individuals to fixed health facilities. Screening with rapid diagnostic test (RDT) followed by parasitological confirmation to be classified as a case. | Self-presentation of symptomatic cases to sub-district health facilities. Screening with RDT followed by clinical confirmation to be classified as a case. | Only about 1% of cases are detected in the US, usually through blood donor screening. Higher levels are detected in endemic areas but this varies widely between different countries. About 6–7 million people are infected worldwide, it is endemic in 21 Latin American countries, but spreading with migrant labour to many other parts of the world [50]. |
| Active/mass screening | WHO guidelines suggest that active screening should be conducted by mobile teams in all villages which have reported cases in the last 3 years and should aim to have 80% coverage. Mobile teams use either the card agglutination test for trypanosomes (CATT) or RDTs for screening before parasitological confirmation of cases. | WHO guidelines recommend that active case detection for VL and PKDL should be conducted routinely, with different intensity approaches employed in different endemicity settings (from a one-off blanket approach in outbreak settings to a year-round index-case-based approach in moderate-to-low endemicity areas) [51]. Individuals are screened for ≥2 weeks fever and hepatosplenomegaly, and suspected cases are tested with an rK39 RDT. | Screening varies widely between different countries. Pregnant women are routinely tested in some South American countries (Argentina, Colombia), but this has been severely impacted by the COVID-19 pandemic. |
| Treatment | Since 2020 the first-line treatment for most cases is fexinidazole (an oral course of treatment), but under 5 year olds, those under 20kg or those with severe symptoms need to take either pentamidine (stage 1 disease) or NECT (stage 2 disease) still. Parasitological confirmation is still required for treatment, however symptoms are not required [28]. | First line treatment for VL is single-dose Liposomal amphotericin B; second-line treatment is daily combination treatment of miltefosine and paromomycin for 10 days. A positive RDT (or biopsy) and clinical symptoms ($\geq 2$ weeks fever and hepatosplenomegaly) are required for treatment [34]. | There have been no new drugs to treat Chagas for 40 years. The antiparasitics, benznidazole and nufurtimox are nearly 100% effective if given to children soon after infection, but only 1% of infected children are ever detected and treated. Up to 40% of infected adults experience adverse reactions: nausea, insomnia, anxiety and depression [46]. |
| Vector control | Vector control is not currently implemented everywhere although it has proved successful at reducing tsetse populations in various gHAT-endemic settings. Primarily this has been done using insecticidal-impregnated "Tiny Targets" deployed along waterways and tsetse population reductions of >80% are typical [15, 44, 29, 26]. | Biannual indoor residual spraying (IRS) of insecticide in all villages that have reported a VL case in the last 3 years is currently implemented [34], despite a lack of evidence of its effectiveness [49]. Long-lasting insecticidal nets have not been used since a randomised controlled trial suggested they have no impact on VL incidence, despite more recent evidence of effectiveness [13]. | Vector control is used at different levels in different locations. This can be intensive within every household to less intensive. Control programmes often only last a few years and the vector populations usually bounce back from surrounding habitats. Intensive vector control can be efficacious when combined with testing and treatment of infants [16]. |

#### Materials and Methods

##### Model description and equations

We use a simple compartmental model to describe the transmission of a vector-borne disease in which asymptomatic infection forms an important part of the natural progression of the disease in terms of duration and transmission potential. The structure of the model is shown in Figure S1 and the corresponding differential equations (1) are given below. We assume an SIR-type (susceptible-infectious-removed) structure for the human population and an SEI (susceptible-exposed-infectious) structure for the vector population. We split asymptomatic infection into non-symptomatic infection $N$ — active infection with the parasite but with no clinical symptoms or progression to them — and pre-symptomatic infection $P$ — active infection, without concurrent symptoms, leading to subsequent clinical symptoms. In the absence of treatment, non-symptomatic individuals either overcome infection and revert to being susceptible (at rate $\tau$) or develop immunity and enter the recovered class $R$ (at rate $\nu$); pre-symptomatic individuals progress to symptomatic infection $I$ (at rate $\varphi$), and symptomatic individuals die (at rate $\alpha$). With existing (or potential) treatments for non-symptomatic, pre-symptomatic and symptomatic infection, individuals are able to recover from each of these infection states (at rates $\gamma_N$, $\gamma_P$ and $\gamma_I$). We include a transmission rate to vectors, $\lambda_V$, which is scaled by a factor dependent on the host's infection status (non-symptomatic, $\varepsilon_N$; pre-symptomatic, $\varepsilon_P$; and symptomatic, $\varepsilon_I$). This is a general scheme for gHAT, VL and Chagas disease. Our assumptions about the definitions of the infection states for each disease are given in Table S2.

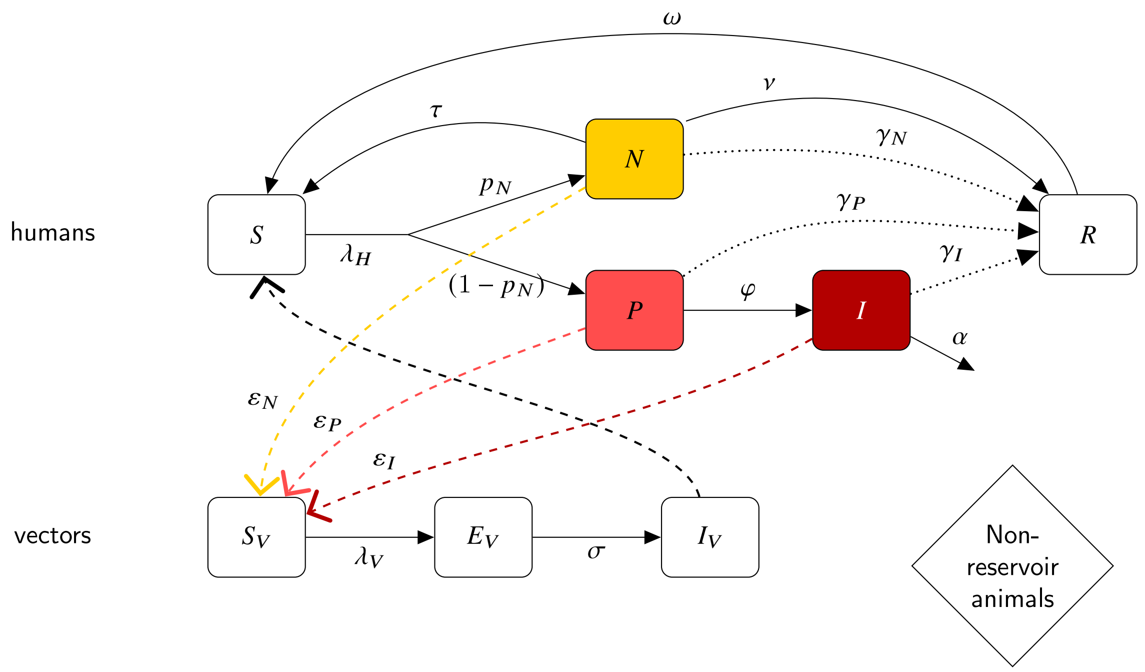


Fig S1: Simple schematic of progression of infection and onwards transmission for a vector-borne disease in which asymptomatic infection is a component. All notation is defined in the *Model description and equations* and Table S3. Parameter values and ranges for each disease we consider (gHAT, VL, Chagas) can be found in Table S3 and Data S1.

$$
\text{Humans}\quad
\begin{cases}
\dfrac{\mathrm{d}S}{\mathrm{d}t} &= \mu_H H + \alpha I + \omega R + \tau N - a p_H f \dfrac{S}{H} I_V - \mu_H S \\
\dfrac{\mathrm{d}N}{\mathrm{d}t} &= p_N a p_H f \dfrac{S}{H} I_V - (\tau + \nu + \gamma_N + \mu_H) N \\
\dfrac{\mathrm{d}P}{\mathrm{d}t} &= (1 - p_N) a p_H f \dfrac{S}{H} I_V - (\varphi + \gamma_P + \mu_H) P \\
\dfrac{\mathrm{d}I}{\mathrm{d}t} &= \varphi P - (\gamma_I + \alpha + \mu_H) I \\
\dfrac{\mathrm{d}R}{\mathrm{d}t} &= (\gamma_N + \nu) N + \gamma_P P + \gamma_I I - (\omega + \mu_H) R
\end{cases}
$$

$$
\text{Vectors}\quad
\begin{cases}
\dfrac{\mathrm{d}S_V}{\mathrm{d}t} &= \mu_V V - a f p_V \dfrac{(\varepsilon_N N + \varepsilon_P P + \varepsilon_I I)}{H} S_V - \mu_V S_V \\
\dfrac{\mathrm{d}E_V}{\mathrm{d}t} &= a f p_V \dfrac{(\varepsilon_N N + \varepsilon_P P + \varepsilon_I I)}{H} S_V - (\sigma + \mu_V) E_V \\
\dfrac{\mathrm{d}I_V}{\mathrm{d}t} &= \sigma_V E_V - \mu_V I_V
\end{cases}
\tag{1}
$$

Table S2: Definitions of asymptomatic (non-symptomatic and pre-symptomatic) and symptomatic infection for gHAT, VL and Chagas.

| | **Asymptomatic** | | **Symptomatic** |
|---|---|---|---|
| **Disease** | **Non-symptomatic** | **Pre-symptomatic** | |
| **Human African trypanosomiasis (gHAT).** Transmitted by tsetse flies, 2,200 cases in 2017, targeted for elimination in 2030. | Parasites detected using parasitological methods for blood/skin, but no development of clinical symptoms. Stage 1 diagnosis likely in active screening. | Same as non-symptomatic except people may have non-specific symptoms (e.g. headache) and will subsequently develop clinical symptoms. Stage 1 diagnosis likely in active screening. | Specific clinical symptoms often promoting health-seeking behaviour such as neurological disturbances associated with stage 2 infection (trypanosomes or elevated white cell count in CSF) |
| **Visceral leishmaniasis (VL)** in the Indian sub-continent[*]. Transmitted by sandflies, 6,700 cases in 2017, targeted for elimination as a public health problem. | Sero-/PCR-positive, e.g. by rK39 RDT/ELISA, DAT or PCR/qPCR, and LST-negative without developing clinical symptoms | Same as non-symptomatic except the individual subsequently develops clinical symptoms | Clinical symptoms incl. prolonged (≥2 wks) fever, hepato-/splenomegaly and rK39 RDT-positive |
| **Chagas disease**[*]. Transmitted by triatomine bugs. Initial infection can lead to symptoms, usually undetected, then progress to chronic indeterminate form | Chronic indeterminate form. Sero-/PCR-positive, by ELISA/IFA/PCR, without symptoms. No ECG abnormalities. | Same as non-symptomatic except subsequently develop clinical symptoms | Sero-/PCR-positive. Chagas cardiomyopathy &/ gastrointestinal disease. ECG abnormalities &/ gastrointestinal dysfunction. |

[*] N.B. The role of post-kala-azar dermal leishmaniasis (PKDL) in transmission of VL in the Indian sub-continent is not considered, and the acute phase of infection in Chagas disease is not modelled here. Pre-symptomatic and symptomatic infections do not precisely align with stage 1 to 2 transition for gHAT, but their duration estimates are used to parameterise the model.

Table S3: Parameter notation and illustrative values used in the analysis in the main text. For full list of parameter ranges see Data S1 supplemental file.

| Notation | Description | gHAT value | VL value | Chagas value |
|---|---|---|---|---|
| $\mu_H$ | Natural human mortality rate | 5.169 $\times 10^{-5}$day$^{-1}$ [19] | 4.551 $\times 10^{-5}$day$^{-1}$ | 4.029$\times 10^{-5}$day$^{-1}$ [47] |
| $\tau$ | Self-reversion ($N \to S$) rate | 0 | 0.00424 day$^{-1}$ [9] (Additional file 1) [5] | 0 |
| $\nu$ | Self-recovery ($N \to R$) rate | 0.001 day$^{-1}$ | 0.00205 day$^{-1}$ [9] (Additional file 1) [5] | 0 |
| $1 - p_N$ | Probability of infection leading to disease | 0.5446 | 0.101 [36] | 0.25 [2, 40, 3] |
| $\varphi$ | $P \to I$ progression rate | 0.0019 day$^{-1}$ [11, 12] | 0.00741 day$^{-1}$ [9] (Additional file 1) | 1.37$\times 10^{-4}$day$^{-1}$ [2] |
| $\alpha$ | disease-induced death rate | 0.0040 day$^{-1}$ [12] | 0.00417 day$^{-1}$ [48] | 2.74$\times 10^{-4}$day$^{-1}$ [45, 18] |
| $\omega$ | Waning immunity rate | 0.0060 day$^{-1}$ [32] | 0 [31, 10] | 0 [3, 40, 35] |
| $H$ | Total human population size | 1000 | 1000 | 1000 |
| $\mu_V$ | Vector mortality rate | 0.03 day$^{-1}$ [41] | 0.0714 day$^{-1}$ [37] | 0.00714 day$^{-1}$ [7] |
| $a$ | Vector bite rate | 0.333 day$^{-1}$ [21] | 0.25 day$^{-1}$ [22, 33] | 0.175 day$^{-1}$ [39] |
| $\sigma$ | Vector incubation rate | 0.04 day$^{-1}$ [41] | 0.143 day$^{-1}$ [23, 37, 42] | 0.125 day$^{-1}$ [38] |
| $f$ | Proportion of blood-meals taken on humans | 1 (reference value) | 1 (reference value) | 1 (reference value) |
| $\varepsilon_N$ | Relative infectiousness of non-symptomatic humans | 0.872 [8] | 0.025 [43, 27] | 1 [20] |
| $\varepsilon_P$ | Relative infectiousness of pre-symptomatic humans | 1 (reference value) | 0.025 (assumed) | 1 [20] |
| $\varepsilon_I$ | Relative infectiousness of symptomatic humans | 0.5 (mid-range value) | 1 (reference value) | 1 [20] |
| $p_V$ | Probability of vector infection from a single bite on an infected human | 1 (reference value) | 1 (reference value) | 1 (reference value) |
| $p_H$ | Probability of human infection from a single bite by an infected vector | 1 (reference value) | 1 (reference value) | 1 (reference value) |
| $m_{eff}$ | Effective vector density | $= p_H V/H$ = 7.20 $\times 10^{-4}$ | $= p_H V/H$ = 0.341 | $= p_H V/H$ = 2.11 $\times 10^{-5}$ |
| $\gamma_N$ | Treatment rate from $N$ | 0 | 0 (no treatment available) | 0 |
| $\gamma_P$ | Treatment rate from $P$ | 0 | 0 (no treatment available) | 0 |
| $\gamma_I$ | Treatment rate from $I$ | 0.002 day$^{-1}$ (assumed) | 0.0111 day$^{-1}$ [6] | 0 |
| $R_c^2$ | Full-cycle reproduction number with basic detection and treatment of symptomatics | 1.1 (mid-range value) | 2.08 (mid-range value) | 1.25* [30] |

* No treatment of symptomatic individuals

#### Calculation of basic reproduction number, $R_0^2$

A key quantity in the control of any infectious disease is the *basic reproduction number*, $R_0$, defined as the average number of secondary infections generated by a single infectious individual in an entirely susceptible population when there are no interventions. If $R_0 > 1$, the disease can survive and spread throughout the population and a stable endemic equilibrium is reached, whereas if $R_0 \leq 1$ the disease will eventually die out. For a vector-borne disease, the parasite is transmitted from one host to another through the vector, so it is often convenient to use the *full-cycle reproduction number*, $R_0^2$, defined as the number of secondary cases in hosts produced by one infectious host in an entirely susceptible population without interventions. The same stability criterion applies to $R_0^2$ as $R_0$ ($R_0^2 > 1$ implies a stable endemic equilibrium, $R_0^2 \leq 1$ implies the disease will go extinct). In practice, vector-borne diseases such as gHAT, VL and Chagas are subject to at least some form of control measures, such as passive detection and treatment of symptomatic cases, so the reproduction number of the disease is calculated taking this into account and termed the (full-cycle) control reproduction number or reproduction number under interventions, $R_c^2$. For the system in (1), $R_c^2$ can be calculated using the next generation matrix approach (see [17]). If vector control leads to a proportional reduction in the vector-to-host ratio of $1 - v$, $R_c^2$ is given by:

$$R_c^2 = \rho(-T\Sigma^{-1})^2, \tag{2}$$

where $T$ is the infection sub-system $(N, P, I, E_V, I_V)$ matrix of transmissions for (1) evaluated at disease-free equilibrium, $(S, N, P, I, R, S_V, E_V, I_V) = (H, 0, 0, 0, 0, V, 0, 0)$:

$$T = \begin{pmatrix} 0 & 0 & 0 & 0 & p_N a p_H f \\ 0 & 0 & 0 & 0 & (1-p_N) a p_H f \\ 0 & 0 & 0 & 0 & 0 \\ v f a p_V \varepsilon_N \frac{V}{H} & v f a p_V \varepsilon_P \frac{V}{H} & v f a p_V \varepsilon_I \frac{V}{H} & 0 & 0 \\ 0 & 0 & 0 & 0 & 0 \end{pmatrix} \tag{3}$$

$\Sigma$ is the matrix of transitions for the same infection sub-system and equilibrium:

$$\Sigma = \begin{pmatrix} -(\tau+v+\gamma_N+\mu_H) & 0 & 0 & 0 & 0 \\ 0 & -(\varphi+\gamma_P+\mu_H) & 0 & 0 & 0 \\ 0 & \varphi & -(\gamma_I+\alpha+\mu_H) & 0 & 0 \\ 0 & 0 & 0 & -(\sigma+\mu_V) & 0 \\ 0 & 0 & 0 & \sigma & -\mu_V \end{pmatrix} \tag{4}$$

and $\rho(-T\Sigma^{-1})$ is the spectral radius of $-T\Sigma^{-1}$, i.e. its largest eigenvalue in absolute value. This yields

$$R_c^2 = \frac{v a^2 f^2 p_H p_V V \sigma}{\mu_V H(\mu_V + \sigma)} \left[ \frac{p_N \varepsilon_N}{(\tau + v + \gamma_N + \mu_H)} + \frac{(1-p_N)\varepsilon_P}{(\varphi + \gamma_P + \mu_H)} + \frac{\varphi}{(\varphi + \gamma_P + \mu_H)} \frac{(1-p_N)\varepsilon_I}{(\alpha + \gamma_I + \mu_H)} \right]. \tag{5}$$

The pre-intervention reproduction number, $R_0^2$, is obtained by setting $v = 1$ (no vector control) and treatment rates to zero in the above expression:

$$R_0^2 \;=\; A \left[ \frac{p_N \varepsilon_N}{(\tau + v + \mu_H)} + \frac{(1-p_N)\varepsilon_P}{(\varphi + \mu_H)} + \frac{\varphi}{(\varphi + \mu_H)} \frac{(1-p_N)\varepsilon_I}{(\alpha + \mu_H)} \right] \tag{6}$$

where $A = \frac{a^2 f^2 p_H p_V V \sigma}{\mu_V H(\mu_V + \sigma)}$. From this expression it can be seen that $R_0^2$ is the product of two major components of the life cycle of each pathogen: a lumped parameter, $A$, that primarily describes parameters associated with the vector (such as the vector-to-host ratio, the bite rate and vector life expectancy), and the sum of the contributions to infection in the vector of each of the human infectious states (non-symptomatic, pre-symptomatic and symptomatic) (Figure S1). The contribution of each infectious state is the product of the time spent in that state, its relative infectiousness to the vector, and the proportion of infections that lead to that type of infectious status.

The proportions of secondary infections arising from non-symptomatics and pre-symptomatics (pre-intervention), $\theta_N$ and $\theta_P$ respectively, are given by:

$$\begin{aligned} \theta_N &= A\left[\frac{p_N\varepsilon_N}{(\tau+\nu+\mu_H)}\right]/R_0^2 \\ \theta_P &= A\left[\frac{(1-p_N)\varepsilon_P}{(\varphi+\mu_H)}\right]/R_0^2. \end{aligned} \tag{7}$$

Let

$$\begin{aligned} x_N &= (\tau+\nu+\mu_H)/(\tau+\nu+\gamma_N+\mu_H), \\ x_P &= (\varphi+\mu_H)/(\varphi+\gamma_P+\mu_H), \\ x_I &= x_P(\alpha+\mu_H)/(\alpha+\gamma_I+\mu_H) \end{aligned} \tag{8}$$

and

$$D = \frac{\varphi}{(\varphi+\mu_H)}\frac{(1-p_N)\varepsilon_I}{(\alpha+\mu_H)}.$$

Then:

$$R_c^2 = \left[\nu x_N\theta_N R_0^2 + \nu x_P\theta_P R_0^2 + \nu x_I AD\right] \tag{9}$$

$$D = \left[R_c^2 - \nu x_N\theta_N R_0^2 - \nu x_P\theta_P R_0^2\right]/\nu A x_I \tag{10}$$

Also

$$\begin{aligned} R_0^2 &= \left[R_0^2\theta_N + R_0^2\theta_P + AD\right] \\ D &= \left[R_0^2 - \theta_N R_0^2 - \theta_P R_0^2\right]/A \end{aligned} \tag{11}$$

So:

$$\begin{aligned} &\left[R_c^2 - \nu x_N\theta_N R_0^2 - \nu x_P\theta_P R_0^2\right]/\nu x_I = R_0^2 - \theta_N R_0^2 - \theta_P R_0^2 \\ &R_0^2\nu(x_I - \theta_N x_I - \theta_P x_I + x_N\theta_N + x_P\theta_P) = R_c^2 \\ &R_0^2 = \frac{R_c^2}{\nu(x_I - \theta_N x_I - \theta_P x_I + x_N\theta_N + x_P\theta_P)} \end{aligned} \tag{12}$$

Rewriting so that $\delta$ is the proportion of treated infections, i.e. $\delta_y = 1 - x_y$ for $y \in \{A, P, I\}$.

$$\begin{aligned} R_0^2 &= \frac{R_c^2}{\nu(1-\delta_I+\theta_N\delta_I+\theta_P\delta_I-\delta_N\theta_N-\delta_P\theta_P)} \\ R_c^2 &= \nu(1-\delta_I+\theta_N\delta_I+\theta_P\delta_I-\delta_N\theta_N-\delta_P\theta_P)R_0^2 \end{aligned} \tag{13}$$

It is useful to note the special case of no vector control ($\nu = 1$) and only basic detection and treatment of symptomatic individuals ($\delta_N = \delta_P = 0$ and $\delta_I = \gamma_I/(\alpha+\mu_H+\gamma_I)$):

$$R_0^2 = \frac{R_c^2}{(1-\delta_I+\delta_I\theta)} \tag{14}$$

where $\theta = \theta_N + \theta_P$ is the total proportion of new infections arising from non-symptomatic and pre-symptomatic infections.

$R_0^2$ can also be written as a function of $R_c^2$ and the contributions from non-symptomatics and pre-symptomatics in the basic treatment scenario, denoted by $\tilde{\theta}_N$ and $\tilde{\theta}_P$ respectively.

Let

$$\begin{aligned} \tilde{\theta}_N &= A\left[\frac{p_N\varepsilon_N}{(\tau+\mu_H)}\right]/R_c^2 \\ \tilde{\theta}_P &= A\left[\frac{(1-p_N)\varepsilon_P}{(\varphi+\mu_H)}\right]/R_c^2 \end{aligned} \tag{15}$$

then:

$$\begin{aligned} \theta_N &= \tilde{\theta}_N R_c^2/R_0^2 \\ \theta_P &= \tilde{\theta}_P R_c^2/R_0^2 \end{aligned} \tag{16}$$

and so

$$\begin{aligned} R_0^2 &= R_c^2/[1-\delta_I+\delta_I(\theta_N+\theta_P)] \\ &= R_c^2/\left[1-\delta_I+\delta_I(\tilde{\theta}_N+\tilde{\theta}_P)R_c^2/R_0^2\right] \end{aligned} \tag{17}$$

$$(1-\delta_I)R_0^2+\delta_I(\tilde{\theta}_N+\tilde{\theta}_P)R_c^2 = R_c^2 \tag{18}$$

$$\begin{aligned} R_0^2 &= \frac{1}{1-\delta_I}\left[1-\delta_I(\tilde{\theta}_N+\tilde{\theta}_P)\right]R_c^2 \\ &= \frac{\alpha+\mu_H+\gamma_I}{\alpha+\mu_H}\left[1-\frac{\gamma_I}{\alpha+\mu_H+\gamma_I}(\tilde{\theta}_N+\tilde{\theta}_P)\right]R_c^2 \\ &= \left[1+\frac{\gamma_I}{\alpha+\mu_H}(1-\theta_c)\right]R_c^2 \end{aligned} \tag{19}$$

where $\theta_c = \tilde{\theta}_N+\tilde{\theta}_P$ is the total proportion of new infections arising from non-symptomatic and pre-symptomatic individuals with basic treatment of symptomatics.

We find the maximum and minimum values of $\theta_c$ across the ranges of the parameter values obtained from the literature (Data S1) numerically (Code_S2) using equation (15) and the expression for $R_c^2$ in (5). We then use equations (16) and (19) to convert the range for $\theta_c$ and the range of literature estimates for $R_c^2$ (Data S1) into ranges for $\theta$ and $R_0^2$.

### Estimated parameters

#### gHAT

##### Estimate of self-recovery rate (from non-symptomatic infection)

We use [24]: Out of 15 people who were originally parasite positive, 9 were parasite negative after around 3.5 years. Assuming exponential decay, with rate parameter $\nu$:

$$\begin{aligned} &\text{Events, } E &&= 9 \\ &\text{Person days, } PD &&= 3.5\times365\times15 \\ &\text{Poisson rate estimate, } \nu &&= \frac{\text{Events}}{\text{Person days}} &&= 4.70\times10^{-4} \text{ per day} \\ &\text{Exact 95\% lower CI} &&= \frac{\chi^2_{2E,0.025}}{2PD} &&= 2.15\times10^{-4} \text{ per day} \\ &\text{Exact 95\% upper CI} &&= \frac{\chi^2_{2(E+1),0.975}}{2PD} &&= 8.92\times10^{-4} \text{ per day} \end{aligned} \tag{20}$$

##### Estimate of waning immunity rate

We use [25]: Out of 39 gHAT patients treated in 1995-1996, Trypanolysis (TL) in 2009 was positive in 24. Assuming that TL positivity is linked to immunity and it has exponential decay, with rate parameter $\omega$:

$$\begin{aligned} &\text{Events, } E &&= 15 \\ &\text{Person days, } PD &&= 13.5\times365\times39 \\ &\text{Poisson rate estimate, } \omega &&= \frac{\text{Events}}{\text{Person days}} &&= 7.81\times10^{-5}\text{per day} \\ &\text{Exact 95\% lower CI} &&= \frac{\chi^2_{2E,0.025}}{PD} &&= 4.37\times10^{-5}\text{per day} \\ &\text{Exact 95\% upper CI} &&= \frac{\chi^2_{(2E+1),0.975}}{PD} &&= 1.29\times10^{-4}\text{per day} \end{aligned} \tag{21}$$

Let $Sk$ be the number of skin-only parasite-positive people in the population and $Bd$ be the blood-parasite-positive people who will not develop disease. We assume that $p_K$ of the infections will lead to skin-only infections, whilst the rest have blood-parasites and $(1 - p_{S|B})$ of these will never have disease and $p_{S|B}$ of these will. I.e. $p_{S|B} = \mathbb{P}(\text{symptoms}|\text{blood infection})$.

The total blood-parasite positive prevalence is $(Bd + P + I)/H$ and the skin-only prevalence is $Sk/H$. Using the following model at equilibrium:

$$\begin{aligned}
\frac{\mathrm{d}Sk}{\mathrm{d}t} &= p_K a p_H f \frac{S}{H} I_V - (\tau + \nu + \gamma_N + \mu_H) Sk_H \\
\frac{\mathrm{d}Bd}{\mathrm{d}t} &= (1 - p_K)(1 - p_{S|B}) a p_H f \frac{S}{H} I_V - (\tau + \nu + \gamma_N + \mu_H) Bd_H \\
\frac{\mathrm{d}P}{\mathrm{d}t} &= (1 - p_K) p_{S|B} a p_H f \frac{S}{H} I_V - (\varphi + \mu_H) P \\
\frac{\mathrm{d}I}{\mathrm{d}t} &= \varphi P - (\alpha + \gamma_I + \mu_H) I
\end{aligned} \tag{22}$$

yields

$$\begin{aligned}
\frac{Sk}{H} &= \frac{p_K a p_H f S^* I_V^*}{H^2(\tau + \nu + \mu_H)} &&= p_K \frac{x}{(\tau + \nu + \mu_H)} \\
\frac{Bd}{H} &= \frac{(1 - p_K)(1 - p_{S|B}) a p_H f S^* I_V^*}{H^2(\tau + \nu + \mu_H)} &&= (1 - p_K)(1 - p_{S|B}) \frac{x}{(\tau + \nu + \mu_H)} \\
\frac{P}{H} &= \frac{(1 - p_K) p_{S|B} a p_H f S^* I_V^*}{H^2(\varphi + \mu_H)} &&= (1 - p_K) p_{S|B} \frac{x}{(\varphi + \mu_H)} \\
\frac{I}{H} &= \frac{\varphi P}{(\alpha + \gamma_I + \mu_H)} &&= (1 - p_K) p_{S|B} \frac{x\varphi}{(\varphi + \mu_H)(\alpha + \gamma_I + \mu_H)}
\end{aligned} \tag{23}$$

If $Sk/H = 0.0054$ and $(Bd + P + I)/H = 0.0175$, then:

$$\begin{aligned}
0.0054 &= Sk/H = p_K \frac{x}{(\tau + \nu + \mu_H)} \\
x &= \frac{0.0054(\tau + \nu + \mu_H)}{p_K}
\end{aligned} \tag{24}$$

$$\begin{aligned}
0.0175 &= (Bd + P + I)/H \\
&= (1 - p_K) x \left[ (1 - p_{S|B}) \frac{1}{(\tau + \nu + \mu_H)} + p_{S|B} \frac{1}{(\varphi + \mu_H)} \left( 1 + \frac{\varphi}{(\alpha + \gamma_I + \mu_H)} \right) \right] \\
&= (1 - p_K) x y
\end{aligned} \tag{25}$$

Solve to find $p_K$ in terms of known (or estimated) values:

$$\begin{aligned}
0.0175 &= (1 - p_K) \frac{0.0054(\tau + \nu + \mu_H)}{p_K} y \\
\frac{0.0175}{0.0054} p_K &= (1 - p_K) y (\tau + \nu + \mu_H) \\
\left( \frac{0.0175}{0.0054} + y(\tau + \nu + \mu_H) \right) p_K &= y(\tau + \nu + \mu_H) \\
p_K &= \frac{y(\tau + \nu + \mu_H)}{\left( \frac{0.0175}{0.0054} + y(\tau + \nu + \mu_H) \right)}
\end{aligned} \tag{26}$$

In [25], out of 53 followed-up patients who refused treatment in 1995/96, by 1999, 9 had become parasite negative, and by 2002 three more were parasite negative. Therefore we might assume that at least 12/53 blood infections result in no symptoms developing (or a maximum of $\hat{p} = 41/53 = 0.7736$ people became symptomatic in the sample, yielding $p_{S|B} < \hat{p} + 1.96\sqrt{\hat{p}(1 - \hat{p})/53} = 0.8863$ as an upper 95% CI bound).

During follow up at least 31 infected people developed symptoms or died due to gHAT disease ($\hat{q} = 31/53 = 0.5849$, yielding $p_{S|B} > \hat{q} + 1.96\sqrt{\hat{q}(1 - \hat{q})/53} = 0.4522$ as a lower 95% CI bound). Other illustrative parameters are taken from the literature (see Data S1) and so (using Code_S2) with a mid-range estimate of $p_{S|B} = 36/53$:

$$\begin{array}{lcl} p_K & = & 0.1982 \\ p_N & = & p_K + (1 - p_K)(1 - p_{S|B}) = 0.4554 \\ 1 - p_N & = & 0.5446 \end{array} \tag{27}$$

## VL

##### Estimate of self-recovery rate (from non-symptomatic infection)

We assume that Leishmanin skin test (LST) positivity is a marker for protective immunity [4] and that "self-recovered" individuals correspond to those who develop LST positivity following non-symptomatic infection (defined as positive serology by rK39 enzyme-linked immunosorbent assay and no clinical symptoms). Let $p_L$ be the proportion who become LST+ following non-symptomatic infection, and $D_N$ be the average duration of non-symptomatic infection. In data from the study of Bern and co-workers in Bangladesh [5], 16 out of 49 non-symptomatically infected individuals became LST+ ($p_L = 16/49 = 0.327$, 95% CI 0.200–0.475), and $D_N = 159$ days (95% CI 138–183 days) from [9] (Additional file 1), so the self-recovery rate $\nu$ can be estimated as:

$$\begin{array}{rcl} \nu = p_L D_N = \frac{16}{49}\frac{1}{159} & = & 0.00205\,\text{day}^{-1} \\ \text{95\% CI: } \left(0.199 \times \frac{1}{183}, 0.475 \times \frac{1}{139}\right) & = & (0.00109, 0.00344)\,\text{day}^{-1} \end{array} \tag{28}$$

##### Estimate of self-reversion rate

Using the same data as above, this can be estimated as:

$$\begin{array}{rcl} \tau = (1 - p_L) D_N = \left(1 - \frac{16}{49}\right)\frac{1}{159} & = & 0.00424\,\text{day}^{-1} \\ \text{95\% CI: } \left((1 - 0.475)\frac{1}{183}, (1 - 0.199)\frac{1}{139}\right) & = & (0.00287, 0.00580)\,\text{day}^{-1} \end{array} \tag{29}$$

#### Additional results

Using the model and disease-specific parameter values (Table S3 and Data S1), we estimate the proportion of new infections arising from asymptomatic individuals for each disease, and use this to assess the likelihood that the diseases can be controlled with different combinations of treatment and vector control interventions, based on published estimates of their basic reproduction numbers (Data S1 and Figure S2). We then consider the dynamic impact of current realistic interventions (Figure 3 in the main text) and potential future alternative interventions (Figures S3–S5) on the prevalence of each of the diseases. In particular, we focus on the relative contribution of non-symptomatic, pre-symptomatic and symptomatic hosts to transmission and how these change over time under different combinations of interventions. All model simulations were performed in MATLAB R2021b [1] and the code is provided in Code_S1.

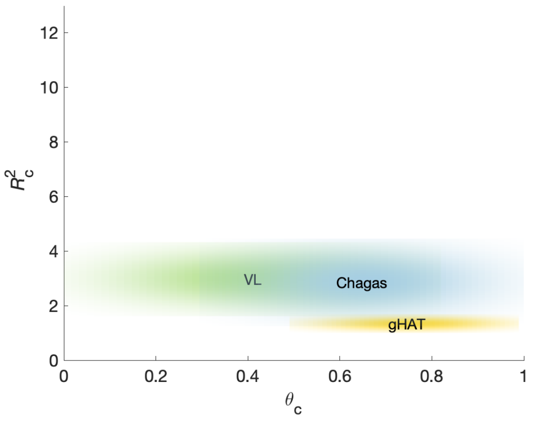


(a)

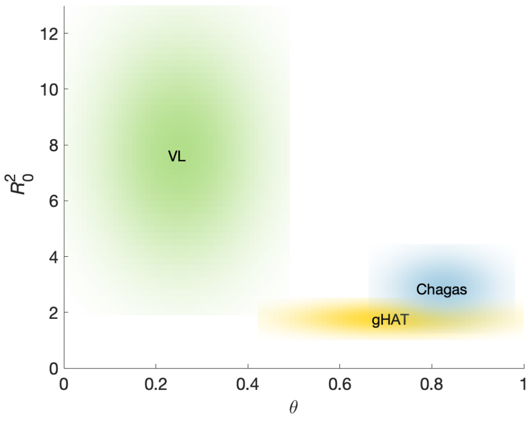


(b)

Fig S2: $R_c^2$ vs $\theta_c$ and $R_0^2$ vs $\theta$ plots. (a) The basic reproduction number, $R_c^2$, for the diseases (gHAT, VL, Chagas) with some basic treatment of symptomatics (see Table S3) compared to the proportion of infections arising from asymptomatics in that setting. (b) The basic reproduction number, $R_0^2$, in the absence of control (epidemiological baseline). Treatment of symptomatics shifts the reproduction number to smaller values ($R_c^2 \leq R_0^2$), and also increases the proportion of cases arising from asymptomatics ($\theta_c \geq \theta$).

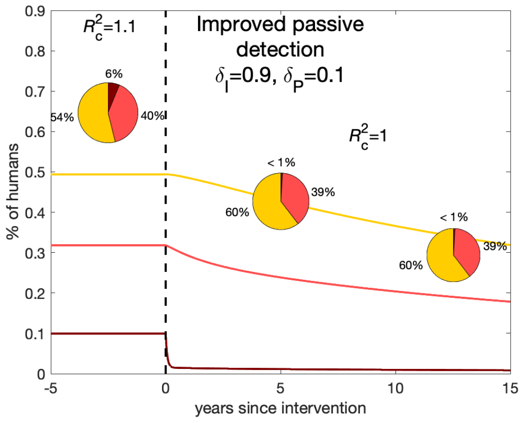


(a)

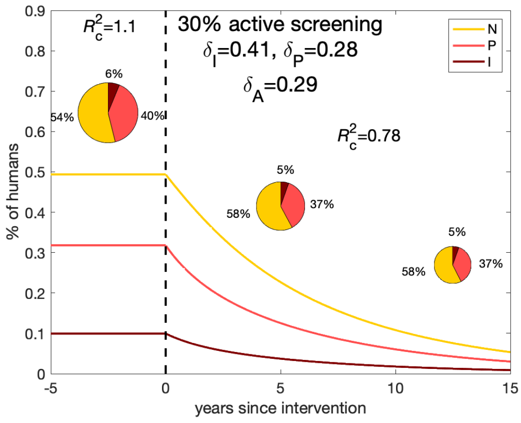


(b)

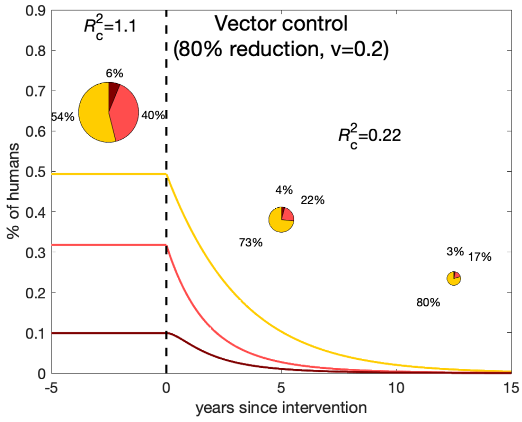


(c)

Fig S3: Impact of different gHAT intervention strategies. Change in contribution of different infection states prior to and throughout an intervention using representative parameters with (a) improved passive detection and treatment so that 90% of symptomatic people and 10% of pre-symptomatic people are treated before death or progression, (b) initiation of active screening so that 30% of the population are screened each year, with the diagnostic algorithm detecting some non-symptomatic infections with blood parasites, and (c) vector control with 80% tsetse population reduction.

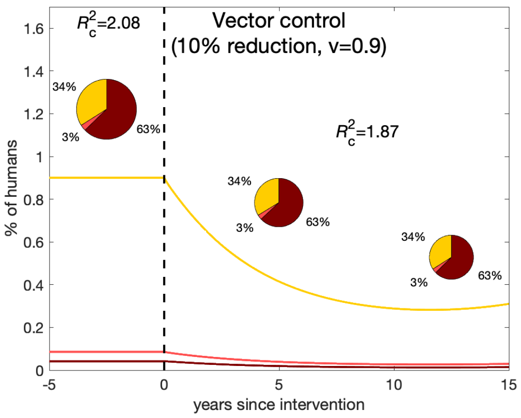


(a)

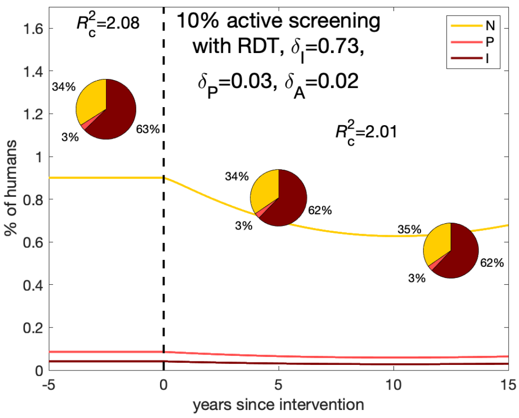


(b)

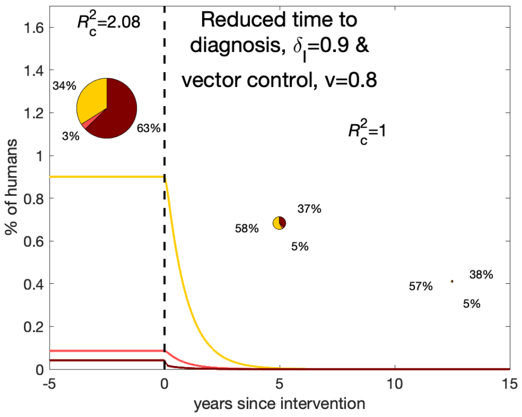


(c)

Fig S4: Impact of different VL intervention strategies. Change in contribution of infected people prior to and throughout an intervention using representative parameters with a) vector control with 10% sandfly population reduction, (b) active screening of 10% of the population in endemic areas with a hypothetical RDT that has sensitivities of 40% and 80% for detecting non-symptomatic and pre-symptomatic infection respectively, and reduces the time to diagnosis for symptomatics such that 90% are treated before death, and (c) a combination of reduced time to diagnosis, such that 80% of symptomatics are treated before death, and vector control with a 30% reduction in the sandfly population.

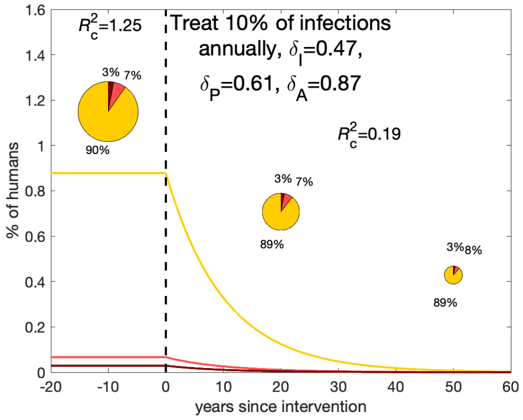


(a)

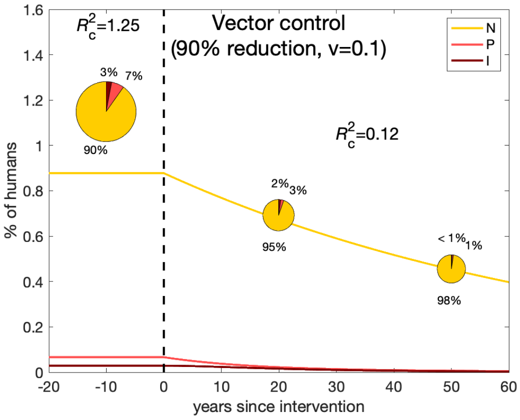


(b)

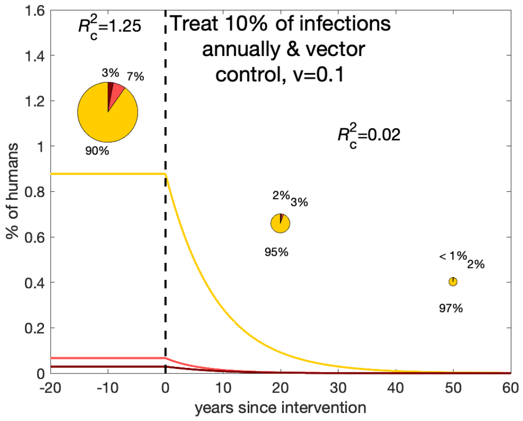


(c)

Fig S5: Impact of different Chagas intervention strategies. Change in contribution of infected people prior to and throughout an intervention using representative parameters with a) treatment of infected individuals such that 10% of infectious people (in each category) are cured each year, (b) vector control with 90% domiciliary triatomine population reduction, and (c) a combination of (a) and (b).

**Data S1.xls** Excel spreadsheet with parameter values and ranges for gHAT, VL and Chagas disease obtained from published literature, and calculated $\theta$ and $R_0^2$ values.

**Code_S1.m** MATLAB code for simulating the transmission model in Figure S1 (system of equations (1).

**Code_S2.m** MATLAB code for finding the minimum and maximum values for $\theta$ and performing the calculations.

[51] World Health Organization. Regional Office for South-East Asia. Process of validation of elimination of kala-azar as a public health problem in South-East Asia, 2016.
